# Fast, efficient and accurate prediction of postoperative outcomes using a small set of intraoperative time series

**DOI:** 10.1101/2024.02.28.24303352

**Authors:** David P. Shorten, Tim Beckingham, Melissa Humphries, Roy Fischer, Natalie Soar, Bill Wilson, Matthew Roughan

## Abstract

In the period immediately following surgery, patients are at high risk of various negative outcomes such as Acute Kidney Injury (AKI) and Myocardial Infarction (MI). Identifying patients at increased risk of developing these complications assists in their prevention and management. During surgery, rich time series data of vital signs and ventilator parameters are collected. This data holds enormous potential for the prediction of postoperative outcomes. There is, however, minimal work exploring this potential. Moreover, existing approaches rely on deep learning, which is computationally expensive, often requiring specialized hardware and significant energy consumption. We demonstrate that it is possible to extract substantial value from intraoperative time series using techniques that are extremely computationally efficient. We used recordings from 66 300 procedures at the Lyell McEwin Hospital (Adelaide, South Australia), occurring in 2013 through 2020. The procedures associated with 80% of the patients were used for model training, with the remainder held out for testing. A combination of techniques including MultiRocket, Multitask and logistic regression were used to predict Rapid Response Team (RRT) calls within 48 hours of surgery and mortality, AKI and elevated troponin levels within 30 days of surgery. This approach achieved an Area Under the Receiver Operating Characteristic curve (AUROC) (95% CI) on the test data of 0.96 (0.95-0.97) for mortality, 0.85 (0.84-0.87) for AKI, 0.89 (0.87-0.91) for elevated troponin levels and 0.80 (0.78-0.83) for RRT calls, outperforming the ASA score and Charlson comorbidity index on the test population for all outcomes. These results show that roughly equivalent accuracy to computationally expensive modelling approaches using diverse sources of clinical data can be achieved using highly computationally efficient techniques and only a small set of automatically recorded intraoperative time series. This implies substantial potential in the analysis of these time series for the improvement of perioperative patient care. We also performed an analysis of the measurement sampling rate required to achieve these results, demonstrating the advantage of high-frequency patient vitals monitoring.

## Introduction

Around 20% of patients experience a significant postoperative complication ^1^ and it has been estimated that at least 4.2 million patients die within 30 days of surgery worldwide each year ^2^. The early identification of patients at risk allows for medical intervention, resulting in risk stratification becoming an important component of anaesthetic evaluation ^3^. As such, there has long been an interest in the derivation of numerical scores indicating a patient’s risk. These have ranged from subjective scores ^4^ to regression models based on demographic variables and surgical parameters ^5–7^. In recent years, there has been increasing interest in the use of machine learning (ML) models for this task ^8–18^. ML models have been able to achieve substantially higher accuracy than traditionally-used scores in the prediction of a number of different postoperative outcomes. In general, they utilize the growing density of electronic data that is collected during a patient’s hospital stay, including demographic data, subjective physician scores, lab test results, patient histories and features of the surgery. A number of ML models also make use of intraoperative variables such as blood loss, medications administered, vitals measurements and ventilator parameters ^9–12^.

Using a large variety of prognostic factors introduces many challenges in the development and deployment of clinical predictive models. The demographic data that is regularly used is often entered manually by healthcare workers with some studies reporting observed error rates of around 3% for manually entered data in clinical settings ^19,20^. These errors will have an impact on the performance of models trained on them ^21,22^. The use of multiple prognostic factors also requires that multiple data types are integrated: demographic data, pathology tests, patient histories among others all need to be associated with a given procedure.

These data are often spread over a patchwork of healthcare systems, never designed to cross communicate or be a platform for prognostic models^13^. These sorts of practical challenges are only compounded when deploying a model in a healthcare system outside of where it was developed. A given procedure or test can be described with different terms across systems and there can be inconsistencies in the meaning associated with a given term ^13,23^. Different systems also collect different data, leading to missing predictor variables at deployment ^24,25^. Even when these variables are available, they might be measured using different procedures, which has been shown to result in model biases ^26^.

In contrast, vitals data and ventilator parameters are routinely collected during surgery by automated systems in a standardized manner ^27,28^. Risk models which are based solely on this data therefore have the potential to be more portable between hospitals and circumvent the requirement for human-recorded inputs.

At present, there exists minimal research exploring postoperative risk models based solely on vitals data and ventilator parameters ^29^. Existing work makes use of 56, disparate surgical time series and creates large deep learning models trained on specialized hardware. In this work, we explore an alternative approach to predicting postoperative outcomes using intraoperative time series. We use a set of 12 time series of routinely observed patient vital signs and ventilator parameters (see Table 2 for a list). We then use a computationally efficient technique, MultiRocket ^18^, to extract around 40 000 random features from the multivariate time series. Multitask lasso regression is used to select features from this larger set, where the ASA score and patient age are used as auxiliary targets. Logistic models are then inferred for each outcome, using the derived feature sets as inputs (See Figure 1 for a diagram of this process). This process requires minimal computational and energy resources; all analysis and model training presented in this paper can easily be run on most modern consumer laptops or PCs.

**Figure 1:**
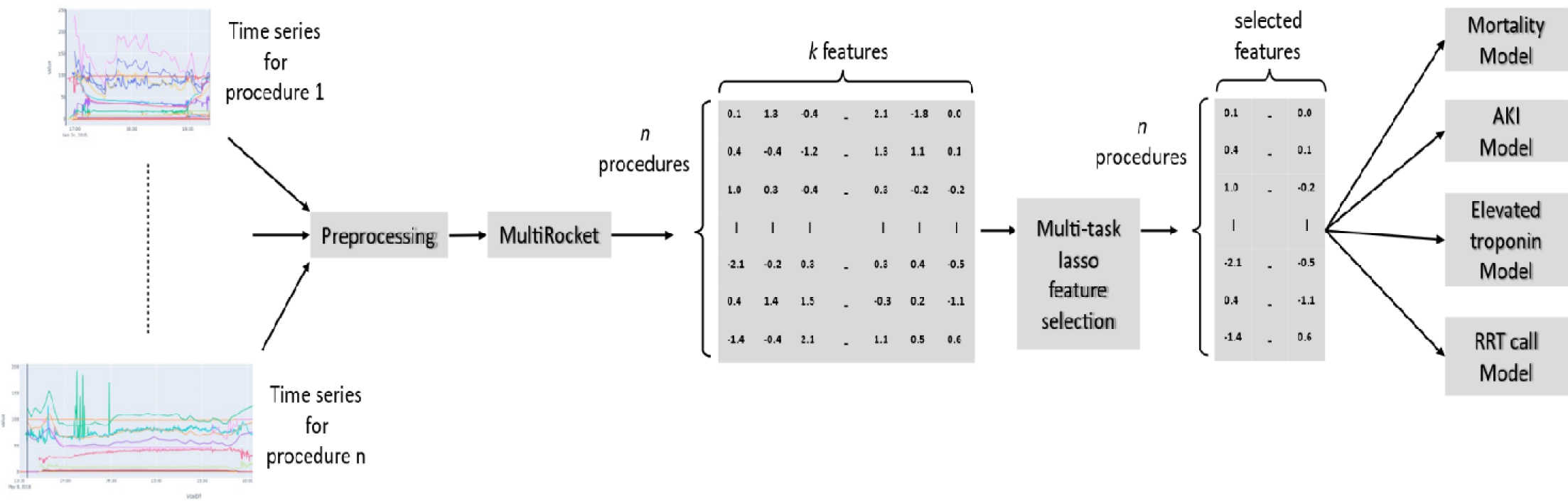
Diagram showing the model fitting procedure.

Despite the limitations in terms of input data and computational resources, the resulting models are effective. On the target outcomes of Acute Kidney Injury (AKI), elevated troponin levels and Rapid Response Team (RRT) calls, the resulting Area Under the Receiver Operating Characteristic curve (AUROC) for the full models is superior to the ASA score ^4^ and Charlson comorbidity index ^30^. Moreover, the resulting AUROC for AKI is similar to that of a recently-published deep learning approach ^29^ which makes use of 56 input channels and requires specialist hardware for training (trained on a different data set, with a similar incidence of AKI: 6.1% vs 5.1% in this study. AUROC 0.81 vs 0.84-0.87 in this study). We also investigated the use of a smaller set of input channels, containing only 4 time series. This was to obtain an indication of how applicable this approach would be to alternate healthcare settings with more limited observations. The performance using this set was within 2% AUROC of that of the full models.

## Results

### Procedure characteristics

Of the 87 765 procedures that occurred at the Lyell McEwin Hospital (Adelaide, South Australia) between January 2013 and November 2020 (inclusive), 66 300 met the inclusion criteria (see Figure 2). Within this selected group, the mean patient age was 47 (std. dev. = 21). The incidence of the outcomes was 441 (0.67%) for mortality, 3365 (5.1%) for AKI, 1129 (1.7%) for elevated troponin levels and 927 (1.4%) for RRT calls. The values of these statistics in the test and training splits, along with additional information, are shown in Table 1. All but four of the used vitals and ventilator channels had recorded data in over 80% of the included procedures. See Table 2 for a full list of the time series channels used and the incidence of absent recordings. Data imputation was not performed in the cases where a recording was not present.

**Figure 2:**
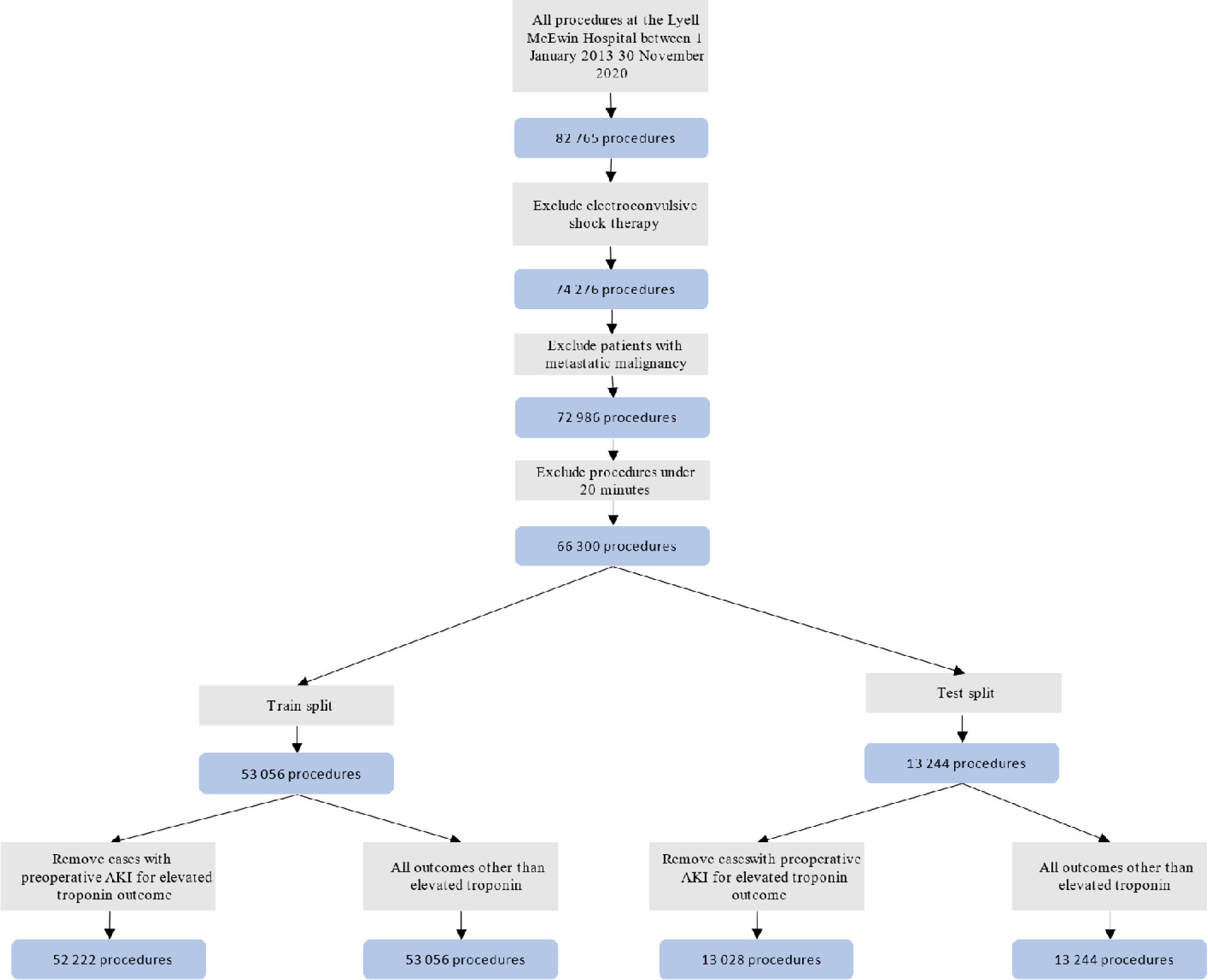
Diagram showing the patient inclusion process.

**Table 1:** Characteristics of the procedures and patients in the training and test sets.

|  | **Training set**<br>**(n = 53,056)** | **Test set**<br>**(n = 13,244)** |
| --- | --- | --- |
| *Patient characteristics* |  |  |
| Number of unique patients | 38,110 | 9,528 |
| Age | 47 s.d. = 21 | 46 s.d. = 21 |
| Preoperative AKI | 834 (1.6%) | 216 (2.4%) |
| ASA score |  |  |
| 1 | 14,635 (28%) | 3,685 (28%) |
| 2 | 24,848 (47%) | 6,158 (46%) |
| 3 | 9,940 (19%) | 2,485 (19%) |
| 4 | 951 (1.8%) | 235 (1.8%) |
| 5 | 46 (0.09%) | 4 (0.03%) |
| unavailable | 2,636 (5.0%) | 677 (5.1%) |
| Charlson comorbidity index |  |  |
| 0 | 27,261 (51%) | 6,925 (52%) |
| 1 | 5,449 (10%) | 1,286 (9.7%) |
| 2 | 5,037 (9.5%) | 1,221 (9.2%) |
| 3 | 4,917 (9.3%) | 1,191 (9.0%) |
| 4 | 4,198 (7.9%) | 1,106 (8.4%) |
| > 4 | 4,312 (8.1%) | 1,038 (7.8%) |
| unavailable | 1,882 (3.5%) | 477 (3.6%) |
| *Procedure Types (MBS<sup>1</sup> code group)* |  |  |
| Group T4: Obstetrics | 8,173 (15%) | 2,190 (17%) |
| Group T8: Surgical operations |  |  |
| 1. General | 16,769 (32%) | 4,077 (31%) |
| 2. Colorectal | 2,681 (5.1%) | 691 (5.2%) |
| 4. Gynaecological | 5,992 (11%) | 1,561 (12%) |
| 5. Urological | 3,811 (7.2%) | 949 (7.2%) |
| 8. Ear, nose and throat | 1,063 (2.0%) | 253 (1.9%) |
| 15. Orthopaedic | 6,988 (13%) | 1,660 (13%) |
| Other surgery | 2,575 (4.9%) | 668 (5%) |
| Other | 328 (0.62%) | 63 (0.48%) |
| Unavailable | 4,676 (8.8%) | 1,132 (8.5%) |
| *Outcomes* |  |  |
| Mortality within 30 days | 351 (0.66%) | 90 (0.68%) |
| AKI within 30 days | 2,741 (5.2%) | 624 (4.7%) |
| Elevated troponin within 30 days | 902 (1.7%) | 227 (1.7%) |
| RRT call within 48 hours | 744 (1.4%) | 183 (1.4%) |

<sup>1</sup> Australian Medicare Benefits Schedule

**Table 2:** List of vital signs and ventilator channels whose time series were used as model inputs. The number of procedures in the test and training sets with records of these channels are shown in the middle columns. The two right-hand columns show their inclusion in the different channel sets.

| Channel | Number of procedures with records |  | 12 channel set | 4 channel set |
| --- | --- | --- | --- | --- |
|  | Training (%)<br>n = 53,056 | Test (%)<br>n = 13,244 |  |  |
| Blood oxygen saturation | 53,018 (>99%) | 13,231 (>99%) | ? | ? |
| Heart rate | 52,822 (>99%) | 13,190 (>99%) | ? | ? |
| Systolic blood pressure | 51,974 (98%) | 13,008 (98%) | ? |  |
| Diastolic blood pressure | 51,974 (98%) | 13,008 (98%) | ? |  |
| Mean blood pressure | 51,961 (98%) | 13,005 (98%) | ? | ? |
| Minimum alveolar concentration | 47,102 (89%) | 11,725 (89%) | ? |  |
| Inspiratory nitrous oxide | 43,104 (81%) | 10,758 (81%) | ? |  |
| Inspiratory oxygen | 43,090 (81%) | 10,752 (81%) | ? |  |
| End-tidal carbon dioxide | 41,506 (78%) | 10,314 (78%) | ? |  |
| Respiration rate | 40,436 (76%) | 10,021 (76%) | ? | ? |
| Inspiratory sevoflurane | 36,688 (69%) | 9,133 (69%) | ? |  |
| Expiratory sevoflurane | 36,688 (69%) | 9,133 (69%) | ? |  |

### Modelling process

The original time series were linearly interpolated at a regular 5 second interval. MultiRocket ^31^ was then applied to the interpolated series to arrive at a set of around 40 000 features. This feature set was reduced using multi-task lasso regression ^32^, where all outcomes were used simultaneously, along with the ASA score and patient age. Separate logistic models were then inferred on the reduced feature set for each outcome (see the Methods section for more details on model development as well as Figure 1). A reduced set of input channels, consisting of a minimal set of 4 channels which contained measurements associated with the minimal standards for anaesthetic monitoring ^33^, was also investigated. Finally, models were also trained on this 4-channel set, but with the interpolation interval increased from 5 seconds to 5 minutes.

The constructed models were evaluated against the ASA score and the Charlson comorbidity index. Missing values of these scores were imputed with the modal value (2 for the ASA score and 0 for the Charlson comorbidity index). Models are evaluated using the Area Under the Receiver Operating Characteristic curve (AUROC). 95% confidence intervals are reported in brackets.

### Full models

The models produced using the full 12-channel input set (and fine, 5-second, interpolation) achieved high accuracy on the test set across all tasks (See Table 3 for a summary of these results. ROC curves for the models are shown in Figure 3). The AUROC scores were 0.96 (0.95-0.97) for mortality, 0.85 (0.84-0.87) for AKI, 0.89 (0.87-0.91) for elevated troponin levels and 0.79 (0.77-0.82) for RRT calls. These scores are significantly higher (by a DeLong test ^34^, p values: 0.007, 4 × 10^-^, 0.002 and 9 × 10^-^, respectively) than the AUROC scores achieved by the ASA score; 0.93 (0.91-0.95) for mortality, 0.82 (0.81-0.83) for AKI, 0.85 (0.83-0.87) for elevated troponin levels and 0.70 (0.67-0.73) for RRT calls. These scores are also significantly higher (p values: 0.007, 0.04, 0.002 and 6 × 10^-^ respectively) than the AUROC scores achieved by the Charlson comorbidity index of 0.92 (0.90-0.94) for mortality, 0.84 (0.82 – 0.85) for AKI, 0.84 (0.81-0.86) for elevated troponin levels and 0.71 (0.68-0.74) for RRT calls.

**Figure 3:**
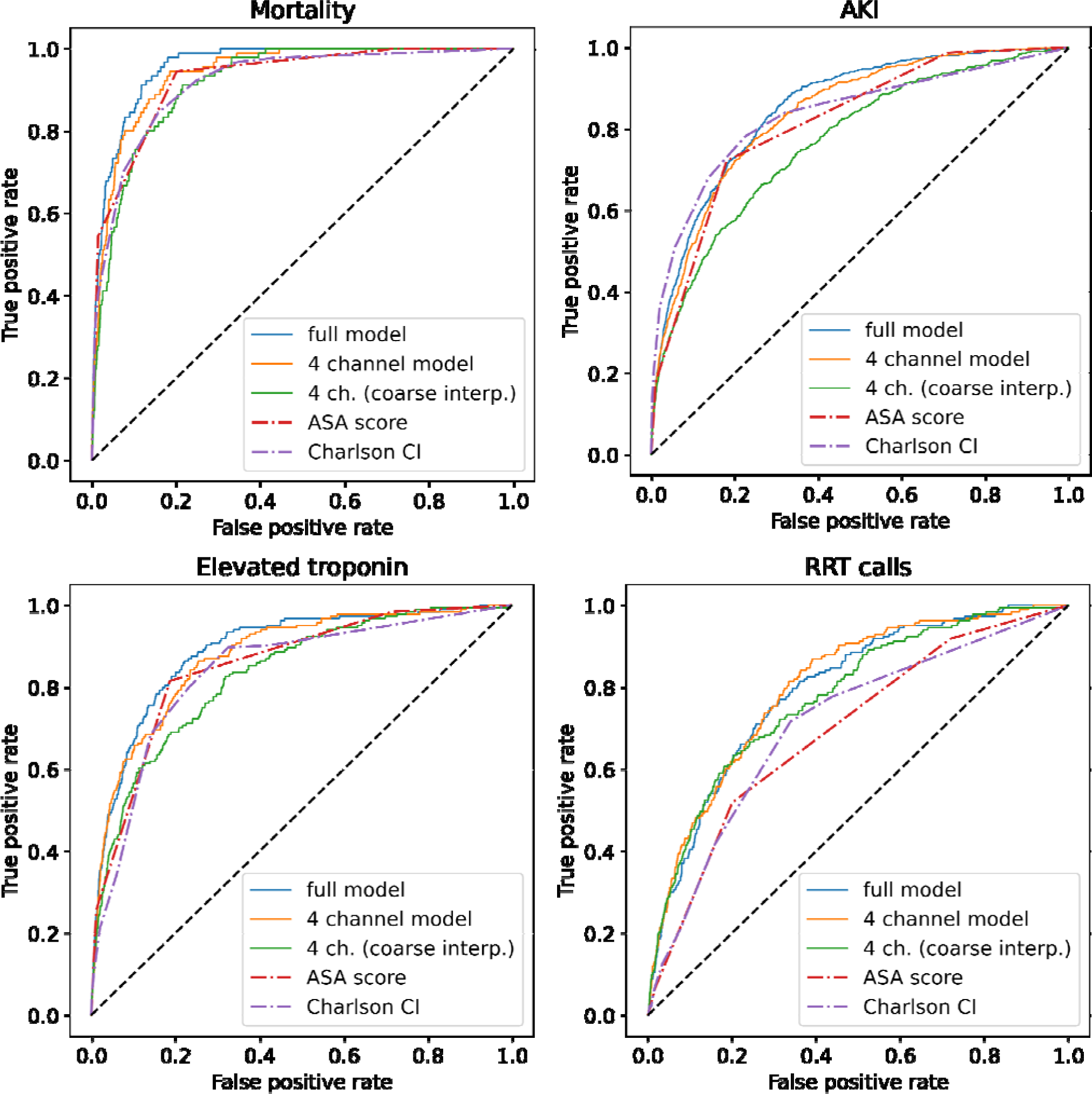
ROC curves for the models on the test set. For the outcomes of mortality, elevated troponin and RRT calls, the full model is able to achieve higher true positive rates across the majority of false positive rates. This difference is substantial for the outcome of RRT calls.

**Table 3:** Accuracy statistics of the models and baseline comparisons (ASA score and Charlson comorbidity index) The models using all 12 input channels consistently achieved the highest area under the ROC curve. These scores dropped slightly when the inputs were changed to the limited 4-channel set. A more substantial drop occurs when the interpolation interval was increased from 5 seconds to 5 minutes.

| **Outcome** | **Prediction** | **AUROC** |
| --- | --- | --- |
| Mortality within 30 days | 12-channel model | 0.96 (0.95-0.97) |
|  | 4-channel model | 0.94 (0.93-0.96) |
|  | 4-channel model (coarse interpolation) | 0.92 (0.90-0.94) |
|  | ASA score | 0.93 (0.91-0.95) |
|  | Charlson comorbidity index | 0.92 (0.90-0.94) |
| AKI within 30 days | 12-channel model | 0.85 (0.84-0.87) |
|  | 4-channel model | 0.83 (0.82-0.84) |
|  | 4-channel model (coarse interpolation) | 0.76 (0.74-0.78) |
|  | ASA score | 0.82 (0.81-0.83) |
|  | Charlson comorbidity index | 0.84 (0.82-0.85) |
| Elevated troponin within 30 days | 12-channel model | 0.89 (0.87-0.91) |
|  | 4-channel model | 0.87 (0.85-0.89) |
|  | 4-channel model (coarse interpolation) | 0.81 (0.79-0.84) |
|  | ASA score | 0.85 (0.83-0.87) |
|  | Charlson comorbidity index | 0.84 (0.81-0.86) |
| RRT call within 48 hours | 12-channel model | 0.80 (0.78-0.83) |
|  | 4-channel model | 0.80 (0.78-0.83) |
|  | 4-channel model (coarse interpolation) | 0.78 (0.75-0.81) |
|  | ASA score | 0.70 (0.67-0.73) |
|  | Charlson comorbidity index | 0.71 (0.68-0.74) |

### Reduced input channel set

Models produced using the smaller set of 4 input channels had AUROC scores of 0.94 (0.93-0.96) for mortality, 0.83 (0.82-0.84) for AKI, 0.87 (0.85-0.89) for elevated troponin levels and 0.80 (0.78-0.83) for RRT calls. Although the differences in score with the full models are small, they are significant (by a DeLong test, p values: 0.02, 3 × 10^-7^, 8 × 10^-4^ and 0.7, respectively) for all outcomes other than RRT calls. The reduced input set models significantly outperformed the ASA score for RRT calls (p = 2 × 10^-8^), but the difference was not significant for mortality, AKI or elevated troponin levels (p values: 0.3, 0.2 and 0.2, respectively). Similarly, it significantly outperformed the ASA score for RRT calls (p = 3 × 10^-7^) but there was no significant difference in the predictive performance for mortality, AKI or elevated troponin levels (p values: 0.2, 0.6 and 0.08, respectively).

### Coarse interpolation

Additional models were trained on the 4-channel input set, but with the interpolation interval increased from 5 seconds to 5 minutes. The resulting AUROC scores were 0.92 (0.90-0.94) for mortality, 0.76 (0.74-0.79) for AKI, 0.81 (0.79-0.84) for elevated troponin levels and 0.78 (0.75-0.81) for RRT calls. Both the 12-channel model and the 4-channel model with fine interpolation significantly outperformed this model on all outcomes (p values: 1X 10^-4^, 4 × 10^-29^, 4 × 10^-9^ and 0.03 between the 12-channel models and coarse sampling and 0.03, 9 × 10^-18^, 1 × 10^-6^ and 0.04 between the 4-channel models and coarse sampling). The coarse interpolation model was significantly outperformed by the ASA score and Charlson comorbidity index for AKI (p values: 1X 10^-7^ and 3 × 10^-10^, respectively), although this relationship was flipped for RRT calls (p values: 6X 10^-5^ and 0.002, respectively). No significant differences were found for mortality (p values: 0.9 and 0.6, respectively). The ASA score significantly outperformed this model for predicting elevated troponin levels (p value: 0.03) but there was no significant difference between its performance and the Charlson comorbidity index (p value: 0.3).

### Comparison of predictions with ASA scores and Charlson comorbidity index

We investigated how closely the predictions of the full model aligned with the ASA scores and Charlson comorbidity index values on the test set. Note that, when performing the following analysis for each score, procedures with missing score values were excluded, whereas they are imputed with the modal values (2 for the ASA score and 0 for the Charlson comorbidity index) in all other parts of this work to allow for their use as auxiliary targets as well as to allow for comparison of predictive ability across the entire test set.

Figure 4 shows the distributions of the predicted probabilities for models produced using all 12 channels for the four outcomes, stratified by the ASA scores of the patients. As the ASA score increased, so did the median of the predicted probabilities for both positive and negative cases. Despite the similarity in trend, positive cases had consistently higher predicted mean/median than negative cases. In particular, for the low ASA score of 2, which captures nearly half of procedures (see Table 1), there is a clear difference in the distributions of the predicted probabilities. Within the subset of procedures with an ASA score of 2, the produced models achieve an AUROC of 0.79 (0.76-0.82) for AKI, 0.83 (0.74-0.91) for elevated troponin levels and 0.78 (0.73-0.82) for RRT calls.

**Figure 4:**
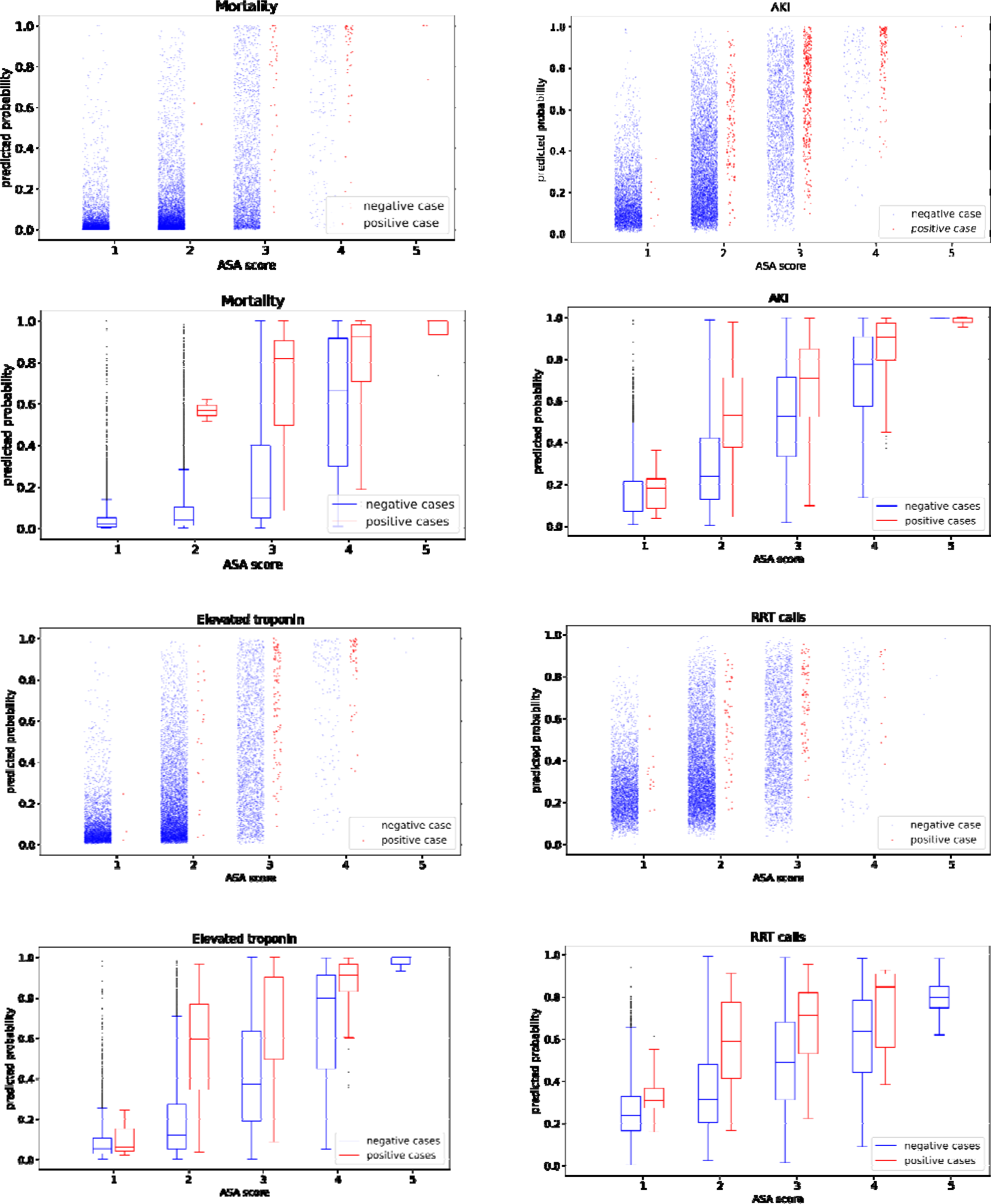
Distributions of predicted probabilities (for the 12-channel models) for outcomes in the test set, stratified by ASA score and outcome. Although the predicted probabilities do increase with the index, the predicted probabilities for positive cases have consistently higher medians than negative cases for a given index value. This indicates that this approach can provide clinicians with information that is complementary to the ASA score.

Figure 5 Shows the distributions of the predicted probabilities for the three outcomes, stratified by the Charlson comorbidity index values of the patients (showing only the common values of four or less). As was the case with the ASA scores, the median of the predicted probabilities increases with the Charlson index. However, positive cases maintained consistently higher predicted probabilities. In particular, for the low Charlson comorbidity index of 0, which captures over half of procedures (see Table 1), there is a clear difference in the distributions of the predicted probabilities for AKI and elevated troponin levels. Within the subset of procedures with a Charlson comorbidity index of 0, the models achieve an AUROC of 0.80 (0.75-0.85) for AKI, 0.83 (0.69-0.95) for elevated troponin levels and 0.68 (0.61-0.75) for RRT calls.

**Figure 5:**
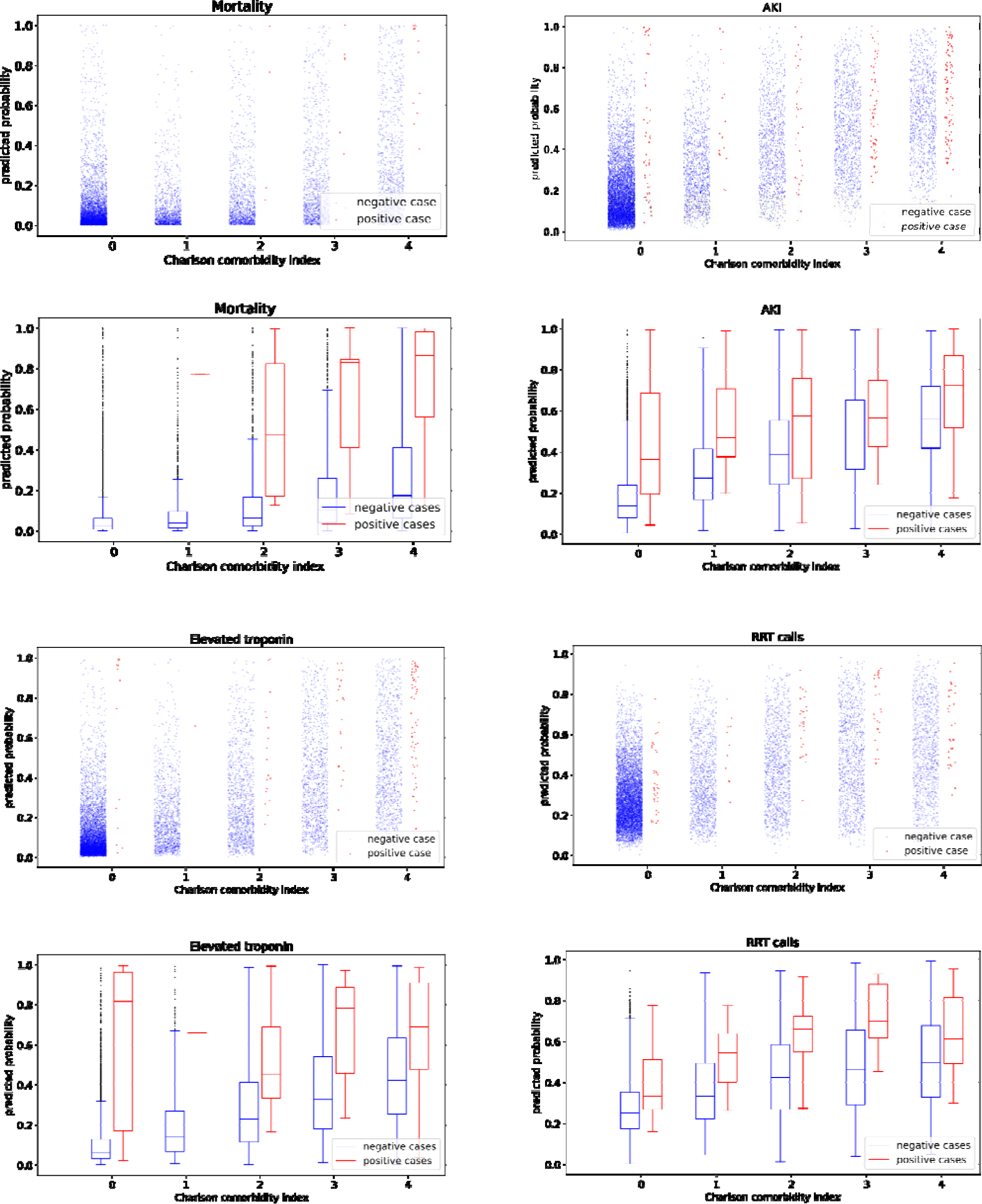
Distributions of predicted probabilities (for the 12-channel models) for outcomes in the test set, stratified by Charlson comorbidity index and outcome. Although the predicted probabilities do increase with the index, the predicted probabilities for positive cases have consistently higher medians than negative cases for a given index value. This indicates that this approach can provide clinicians with information that is complementary to the Charlson comorbidity index.

It was not possible to perform a similar analysis for the modal ASA score and Charlson comorbidity index values for the mortality outcome, due to the low numbers of positive cases associated with these scores. For cases with an ASA score of three, models trained on all 12 channels achieved an AUROC of 0.82 (0.77-0.86). For cases with a Charlson comorbidity index of 3, the full models achieved an AUROC of 0.80 (0.72 – 0.88).

## Discussion

We have demonstrated that a simple, computationally efficient modelling framework based only on a small set of intraoperative time series can achieve similar accuracy to large deep learning models based on wide feature sets. We developed a machine-learning pipeline which performs postoperative outcome prediction using only data from intraoperative time series. The pipeline first extracts features using an efficient technique, MultiRocket ^31^. A smaller subset of features is then selected using multi-task lasso ^32^, where both age and ASA score are used as auxiliary targets. Finally, separate models for the different outcomes (mortality, AKI, elevated troponin levels and RRT calls) are trained on the reduced feature sets. On the outcomes of elevated troponin levels and RRT calls, this modelling approach achieved performance scores significantly higher than the commonly-used ASA score or the Charlson comorbidity index. Moreover, predicted scores were similar to those achieved by top-performing models which make use of much larger and less reliable input sets ^9,11,12,15^. Further, this approach was able to achieve an AUROC for AKI equivalent to a large deep learning model trained using specialized hardware on a much larger set of 56 intraoperative time series ^29^ (trained on a different data set, with a similar incidence of AKI: 6.1% vs 5.1% in this study. AUROC 0.81 vs 0.84-0.87 in this study). These results indicate the substantial predictive value that can be found in intraoperative time series.

There are multiple benefits to our proposed approach. The first group of these benefits derive from the models only making use of a small set of intraoperative time series from vitals monitors and ventilators. Relying on data that is entered by humans increases the chances of input error or missing values. Further, some models are limited to patients that have preoperative blood tests^8–10,14,16^ or use procedural data normally collected at patient discharge^6,7,11,15^, making it challenging to predict prospectively. Moreover, there is variation in how certain clinical quantities are recorded and measured across different locations, which can result in model biases ^26^. By contrast, there are standards and established guidelines concerning which vitals and ventilator parameters need to be observed during surgery ^33,35^ and data are collected automatically by standardized devices ^27,28^. Given that the models do not require the ventilator and vitals data to be integrated with other data sources, they can be deployed at the point of data collection. This indicates that they have the potential to be deployed with comparative ease across multiple healthcare centers. It is hoped that these features will facilitate future prognostic validation in clinical practice.

The second benefit is one of computational efficiency and accessibility. The dominant technique in recently-developed post-operative risk models is deep learning ^3,9–11,13,14,16,17,29^. Although deep learning is capable of producing highly-performant models, this comes at the price of high computational expense, often entailing specialized hardware and significant energy expenditure ^36^. The models used here make use of an extremely computationally efficient technique, MultiRocket ^31^, allowing them to be trained quickly on most modern consumer hardware. MultiRocket was able to train a single resample of 112 UCR benchmark^37^ time series classification problems in under 16 minutes^31^. Deep learning methods of comparable performance took days or weeks on the same task^31^. Not only does this computational efficiency make this approach more accessible to diverse practitioners, but it facilitates rapid prototyping on new datasets. State of the art deep learning hardware such as the Nvidia H100 cost in the region of USD 30,000 with limited availability^38^. Given that patient data is owned by disparate organizations worldwide, demonstrating that value can be obtained from this data using consumer-grade hardware will hopefully motivate these organizations to take full advantage of it.

Thirdly, we have shown that models trained on a limited set of channels that are close to minimum anaesthetic monitoring standards ^33,35^ produced similar accuracy to the larger models. This is a positive indication of the potential for this approach to produce models which can be deployed in settings with more limited access to patient measurements.

The sampling rate of vitals recordings varies greatly across healthcare settings. As such, it is useful to analyze the impact of lower interpolation rates (which would be impervious to lower raw sampling rates) to our modelling approach. We trained models on the 4-channel input set with the interpolation interval increased from 5 seconds to 5 minutes. This combination of input channels and sampling rate approximates the data available in ICU settings in the local hospital system. It was found that this large increase in the interpolation interval resulted in a substantial decrease in the performance of the models. This indicates that deployment of this approach is likely to require reasonable data sampling rates. Future work will attempt to further elucidate the sampling rate requirements and techniques for mitigating this drop in performance.

We were also interested in ascertaining whether there was potential for this framework to provide clinically useful information beyond the ASA scores and Charlson comorbidity index. We achieved this by investigating the distributions of model predictions at different patient ASA scores and Charlson comorbidity index values (the results are plotted in Figures 2 and 3 and discussed in Results). While the model predictions did not differ substantially between positive and negative cases for high ASA scores, there are large differences in the distributions of the predictions for the lower scores. Nearly half of patients in this dataset had an ASA score of 2 (see Table 1). This is a large pool of patients who are considered low risk. Identifying the patients within this pool who have a high risk of developing complications could be clinically useful diagnostically but also as a screening tool. Our models showed strong discriminative ability within this group of patients, indicating their ability to flag at-risk patients who might otherwise have been overlooked. Similarly, over half of patients in the procedures included in this study received a Charlson comorbidity index of 0, indicating that they are considered low risk by this measure. For the outcomes of elevated troponin and AKI, our models demonstrated strong discriminative ability within this pool.

It is worth briefly noting that the ASA scores in this dataset were lower than in some similar studies^8–10^, where the modal score was 3 compared to 2 here (see Table 1). However, other studies ^7,16^ had ASA scores closer to those in this dataset. Similarly, over half of the patients in our study received a Charlson comorbidity index of 0.

A limitation of the presented approach, and many ML approaches, is that it does not provide for easy interpretability. In the context of medical decision making, there is substantial value in models which provide interpretable answers ^18,39,40^. Not only does this increase physician trust in the model but discovered markers of adverse outcomes can be taken as the starting point for further study. They provide insight into the possible underlying mechanisms that may give rise to the physical outcomes we observe. Methods for interpreting black-box models, such as ours, do exist ^41,42^, and we intend to apply them to our models in future work. However, it is unlikely that the level of interpretability achieved will approach an intrinsically interpretable modelling technique.

Following from this, models trained and evaluated on one center are not guaranteed to achieve high performance on data from another center ^43,44^. This was a single-centered study and did not include patients that underwent specialist surgery such as cardiothoracic or neurosurgery. An important component of future work will be the evaluation of these models on externally obtained data containing a wider variety of procedures.

In terms of the data used to train the model, future work may consider incorporating data on patient urine output in order to evaluate the urine output criterium of the KDIGO criteria for stage 1 AKI ^45^. This data was not available for the current study, so some proportion of the patients in both the training and test sets of our data who were encoded as not having an AKI diagnosis may have met the KDIGO criteria by this third criterium. This is a common limitation in machine learning models for the prediction of AKI ^40,46–48^ and would be worth addressing if designing future data collection.

Another limitation of this study for the outcomes of troponin elevation and AKI (determined via a test for creatinine elevation) is that the data does not necessarily contain all patients that had an elevation. Some patients in the dataset did not receive blood tests for these markers, and so the true rate of these outcomes might be higher than what was observed. Our rate of AKI (∼5%, see Table 1), was lower than that reported in other studies (6.% to 15% ^10–12,14^). The lack of comprehensive screening might contribute towards this low rate.

A final limitation, specific to the outcome of RRT calls, is that the structure of these calls changed at the Lyell McEwin Hospital in early 2020, transitioning from a single to a two-tiered system. The creation of a second tier changed the threshold for MER calls following its introduction, potentially influencing the accuracy of the model.

In summary, we have developed a simple, computationally efficient model based only on a small set of intraoperative time series that predicts postoperative outcomes with reliability similar to large, computationally expensive alternatives. Our modelling framework is fast, accessible and can discriminate cases that will develop postoperative complications from those that will not. Although the generalizability of this work should be tested, it shows great promise for a reliable way of predicting outcomes, even with sparse data. We hope that this will motivate further research into the use of these time series for postoperative outcome prediction.

## Methods

This was a single center retrospective study used to develop a prognostic model. The TRIPOD guidelines^49^ have been followed for reporting. Ethical approval was granted by Central Adelaide Local Health Network Human Research Ethics Committee (reference number HREC16298). Local governance approval from the Northern Adelaide Local Health Network was also obtained (SSA 22 033).

### Data source

Data for patients at the Lyell McEwin Hospital, Adelaide, was collected retrospectively from the South Australian Health electronic medical record system and directly from the electronic anaesthetic information management system WinChart. We included all surgical procedures at this hospital occurring between January 2013 and November 2020 (inclusive), subject to the following exclusions. All electroconvulsive shock therapy, procedures lasting less than 20 minutes, procedures where the patient was under the age of 18 and procedures where the patient was diagnosed with metastatic cancer were excluded. For the elevated troponin target, all procedures where kidney failure was diagnosed at the time of the procedure were excluded (this was only done when training the final models, not for feature selection). See Figure 2 for a diagram of this exclusion process. Some patients underwent multiple procedures during the time period of this study. Care was taken to ensure that for all train-test and cross-validation splits, all procedures associated with a given patient were assigned to one side of the split.

### Target and score determinations

Targets were determined based on data in patient electronic health records. Age, ASA score, mortality and RRT calls were associated with cases in a straightforward manner. AKI and elevated troponin levels were inferred from pathology records (described below). AKI and elevated troponin were observed within a 30 day window and RRT calls were observed within a 48 hour window after the completion of the procedure. Missing values were imputed for the ASA score and age. Missing ASA scores were assigned the modal value of two. Note that this was done both for its use in the multi-task feature selection and as a baseline prediction against which to compare the performance of derived models. Missing age values were assigned the median age. The Charlson comorbidity index was determined from ICD-10 codes, using an existing, validated procedure ^30^. Missing Charlson comorbidity index values were assigned the modal value of zero.

### AKI

AKI was determined from creatinine tests according to the KDIGO definition ^45^. This process was performed using the tests from the Lyell McEwin hospital as well as other hospitals within South Australia, to detect AKI in patients who were transferred after surgery. We detected AKI based on either a raise in serum creatinine measurements over time or a single measurement of serum creatinine above a threshold. A raise was detected if a given measurement was at least 26.5 µmol/l greater than a previous measurement taken within the preceding 6 months. A single measurement of creatinine was taken to imply AKI if it was greater than 1.5 time baseline.

#### Elevated troponin levels

Troponin levels were interpreted as elevated if they were greater than the 99^th^ percentile of these measurements in the local system (29 ng/L).

### Time series preprocessing

The original time series were irregularly sampled, with differences in the average inter-sample interval between channels. Channels also had different start and end points. This necessitated significant preprocessing. No outlier removal was performed.

#### Start and end determination

For each procedure, the different channels within the multivariate time series had slightly different start and end points. This necessitated the determination of common start and end points of each procedure. The start point was chosen to be the time point by which 40% of the vitals and ventilator channels (within the 12-channel set, see Table 2) which had any recorded values had had their first recorded value. The end of the procedure was determined by a sliding window approach, with a window of length 30 seconds being moved across the procedure with a step size of 10 seconds. When the total number of sample points within the window dropped below 10% of the median seen in previous steps, the procedure was taken to have ended at the start of the time window (with at least 10 window shifts having already been observed).

#### Interpolation

Some of the vitals and ventilator values were sampled irregularly. Moreover, they had different sampling rates. In order to produce a regularly sampled multivariate time series, each channel was linearly interpolated at an interval of either 5 seconds (for the regular models) or 5 minutes (for the low interpolation rate models). Before the first and after the last samples of a given channel, the values were backfilled or forward filled. That is, before the first sample, the value of the first sample was used at all time points. Similarly, after the last sample, the value of the last sample was used at all time points.

#### Missing channels

Some vitals and ventilator channels were not recorded in certain procedures (see Table 2 for a list of the incidence of missing recordings). The values for missing recordings were not imputed. They were coded as zero before feature extraction by MultiRocket.

#### Scaling

After interpolation, each recorded channel was scaled to have zero mean and unit variance, calculated across all recordings. This was performed prior to feature extraction by MultiRocket. MultiRocket draws kernel weights from a predetermined limited set of 84 weight combinations ^31^. Channels having different scales would have the equivalent effect to modifying the kernel weights and biases, hence the need for scaling prior to feature extraction.

#### Respiration rate in the presence of paralytics

As patients administered paralytics will have a constant respiration rate determined by the ventilator, in cases where paralytic medication was administered, the respiration rate channel was treated as missing even if it was recorded. The specific paralytics present in this procedure pool were Rocuronium and Vecuronium.

### Model construction

#### Feature Extraction

Features were extracted using MultiRocket ^31^, which uses semi-random kernels to extract a large bank of features against which regression can be performed. Small modifications were made to the original algorithm. Maximum pooling was added to the set of pooling features and the mean index pooling was excluded. Preliminary testing indicated that this improved performance on this task. Instead of performing the feature extraction separately on both the raw time series and their differences, we combined these into a single multivariate time series and performed extraction on them jointly. This was indicated to be effective by preliminary testing.

The bias fitting procedure was also changed to allow for the variable length time series in this study. The length parameter required in this fitting procedure was set as the median length across the training set. The rest of the fitting procedure proceeded using only the recordings of length greater than the median.

All features were scaled to have zero mean and unit variance after extraction.

To aid in reproducibility, the modified version of MultiRocket code used in this study is provided with the rest of the analysis code which will be made available on publication.

#### Feature selection

A subset of features was selected from the original set generated by MultiRocket. This was performed using multitask lasso regression, where the weights were penalized using the *l*_2,1_ norm. This is an established technique for feature selection where multiple targets share similar relevant features ^32,50,51^. We optimize

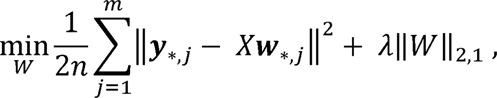

Where

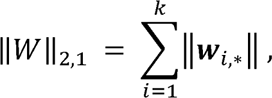

*n* is the sample size, m the number of targets, *k* the original number of features and λ is the regression penalty. The matrix *Y* is the *n* x *m* target matrix, *X* is the *n* x *k* design matrix and *W* is the *k* x *m* weight matrix. The notation ***w****_*,j_* is used for a column of W and ***w***_i,*_ denotes a row. The lasso penalty across the rows will encourage each entire row to be set to zero. We interpret a feature as selected if there are any non-zero elements in the corresponding row of W.

The multitask target used for feature selection was set as all targets in the current study, augmented with the patient age and ASA score. Using these additional targets to assist in the feature selection was found to yield improved model performance in the downstream regression tasks, particularly for targets with a lower incidence of positive outcomes. The binary outcomes were integer encoded, before all outcomes were scaled to have zero mean and unit variance.

#### Final outcome models

Separate logistic ridge regression models with balanced class weighting ^52^ were inferred for each outcome, based on the set of features selected by multitask lasso.

#### Penalty selection

Separate feature selection lasso penalties λ_i_ and corresponding final model ridge penalties α_i_ were selected for each outcome independently, using nested cross validation on the training set. For each outcome;, a set of lasso regression penalties Λ_i_ were evaluated on each of 4 cross-validation folds. For each cross-validation fold, for each λ_i,j_ E Λ_*i*_, multitask lasso feature selection was performed with that penalty to select a set of features *F_i,j_* using the fold’s training set. 4-fold cross validation was then run using this selected feature set **F_i,j_**, in order to select an optimal ridge regression penalty α_*i,j*_ from a set of candidate penalties Α_i_. The average precision on the fold’s test set, for the model inferred with the optimal penalty α_*i,j*_ was then used as the performance metric associated with the tested λ_i,j_ on this particular fold. An optimal λ_i,*_ was then chosen as the λ_i,j_ with the highest mean of the mean precision on the validation set across all folds. The final model for outcome; was then trained on the entire training set, where feature selection was first performed using the chosen λ_i_ before an associated α_*i*_ was chosen with four-fold cross validation on the entire training set. For efficiency reasons, we used a common set of regression penalties Λ for all outcomes. This allowed for the feature selection to be pre-computed across all training sets of the 4 folds, and then re-used for each outcome. A common set of ridge regression penalties A was also used both within cross validation and for the final model of each outcome. For Λ, we used 5 linearly spaced values between 0.02 and 0.05 (inclusive). For A, we used 15 logarithmically spaced values between 1×10-^5^and 1×10^2^ (inclusive).

#### Model testing

Models were evaluated on a test set of 8913 procedures. This test set was chosen by selecting all procedure associated with 20% of patients (chosen randomly). Confidence intervals of the AUROC scores were estimated using 1000 bootstrap resamplings.

## Code availability

The code for training the models and producing the figures in this text will be made available on publication.

## Data Availability

The data used in this study contains private patient information and so will not be made available to other parties.

## Acknowledgements

This work was funded by a University of Adelaide ECMS Strategic Funding grant. This research was conducted using data provided by the Northern Adelaide Local Health Network (NALHN). Many staff members at NALHN provided valuable advice and assistance at all stages of this research.

